# Long-term symptoms after SARS-CoV-2 infection in school children: population-based cohort with 6-months follow-up

**DOI:** 10.1101/2021.05.16.21257255

**Authors:** Thomas Radtke, Agne Ulyte, Milo A Puhan, Susi Kriemler

## Abstract

Although long COVID in children exists, it is still unclear to what extent children are affected. The *Ciao Corona* study is a longitudinal cohort investigating SARS-CoV-2 seroprevalence and clustering of cases among around 2500 children and adolescents (hereafter referred to as children) from 55 randomly selected primary and secondary schools in the canton of Zurich in Switzerland. Between June 2020 and April 2021, we completed three testing phases where we collected venous blood for serological analysis and asked about symptoms with online questionnaires. We compared children who tested positive for SARS-CoV-2 antibodies in October/November 2020 with those who tested negative. Children who were seronegative in October/November 2020 and seroconverted or were not retested in March/April 2021 were excluded from the analysis (n=256). In March-May 2021 we assessed the presence of symptoms occurring since October 2020, lasting for at least 4 weeks, and persisting for either >4 weeks or >12 weeks. Overall, 1355 of 2503 children with a serology result in October/November 2020 and follow up questionnaire in March-May 2021 were included. Among seropositive and seronegative 6-to 16-year-old children, 9% *versus* 10% reported at least one symptom beyond 4 weeks, and 4% *versus* 2% at least one symptom beyond 12 weeks. None of the seropositive children reported hospitalization after October 2020. This study suggests a low prevalence of symptoms compatible with long COVID in a randomly selected population-based cohort of children followed over 6 months after serological testing.

## INTRODUCTION

Children can suffer from SARS-CoV-2 postviral syndromes, but it is yet unclear to what extent children are affected by long COVID, herein defined as symptoms that continue for more than 12 weeks after SARS-CoV-2 infection and are not explained by an alternative diagnosis.^1^ Current evidence is predominantly limited to selective – mostly clinical – populations without control groups^2–5^, which do not allow estimating the overall prevalence and burden in a general pediatric population. In this study, we compared long COVID compatible symptoms in children with 6-months follow-up according to their SARS-CoV-2 serology.

## METHODS

The *Ciao Corona* study is a longitudinal cohort investigating SARS-CoV-2 seroprevalence and clustering of cases among around 2500 children from 55 randomly selected primary and secondary schools in the canton of Zurich (about 1.5 million inhabitants) in Switzerland (study design described elsewhere^6^). Between June 2020 and April 2021, we completed three testing phases where we collected venous blood for serological analysis and asked about symptoms with online questionnaires. For serological analysis, we used the ABCORA 2.0 test.^6^

In this analysis, we compared children who tested positive for SARS-CoV-2 antibodies in October/November 2020 with those who tested negative. Children who were seronegative in October/November 2020 and seroconverted or were not retested in March/April 2021 were excluded from the analysis (n=256). In March-May 2021 we assessed the presence of symptoms occurring since October 2020 and lasting for at least 4 weeks^3^, and persisting for either >4 weeks or >12 weeks.

Descriptive analysis was performed with R version 4.0.3. The Ethics Committee of the Canton of Zurich, Switzerland (2020-01336) approved the study and parents/legal guardians provided written informed consent.

## RESULTS

Overall, 1355 of 2503 children with a serology result in October/November 2020 and follow up questionnaire in March/April 2021 were included. Participant characteristics, symptoms and self-rated health are summarized in the Table. Among seropositive and seronegative children, 9.2% (10/109) *versus* 9.7% (121/1246) reported at least one symptom beyond 4 weeks, and 3.7% (4/109) *versus* 2.2% (28/1246) at least one symptom beyond 12 weeks (Table). None of the seropositive children reported hospitalization after October 2020. The distribution of pre-existing chronic health conditions among seropositive (n=109) and seronegative children (n=1246) is given in Supplemental Table S1. Participant characteristics, symptoms and self-rated health for a subpopulation of seropositive (n=89) and seronegative children (n=891) without chronic health conditions is shown in Table S2.

**Table.**
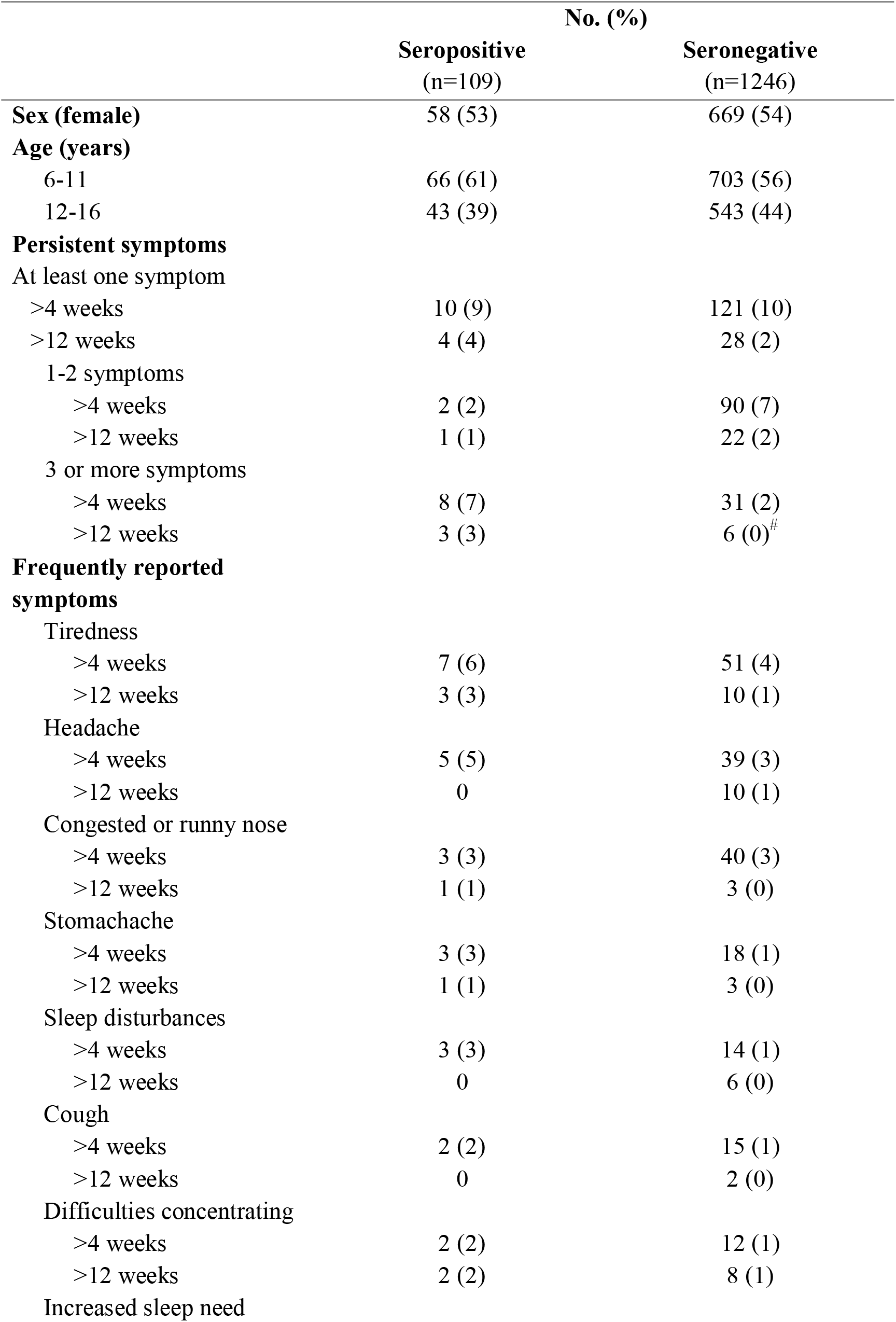
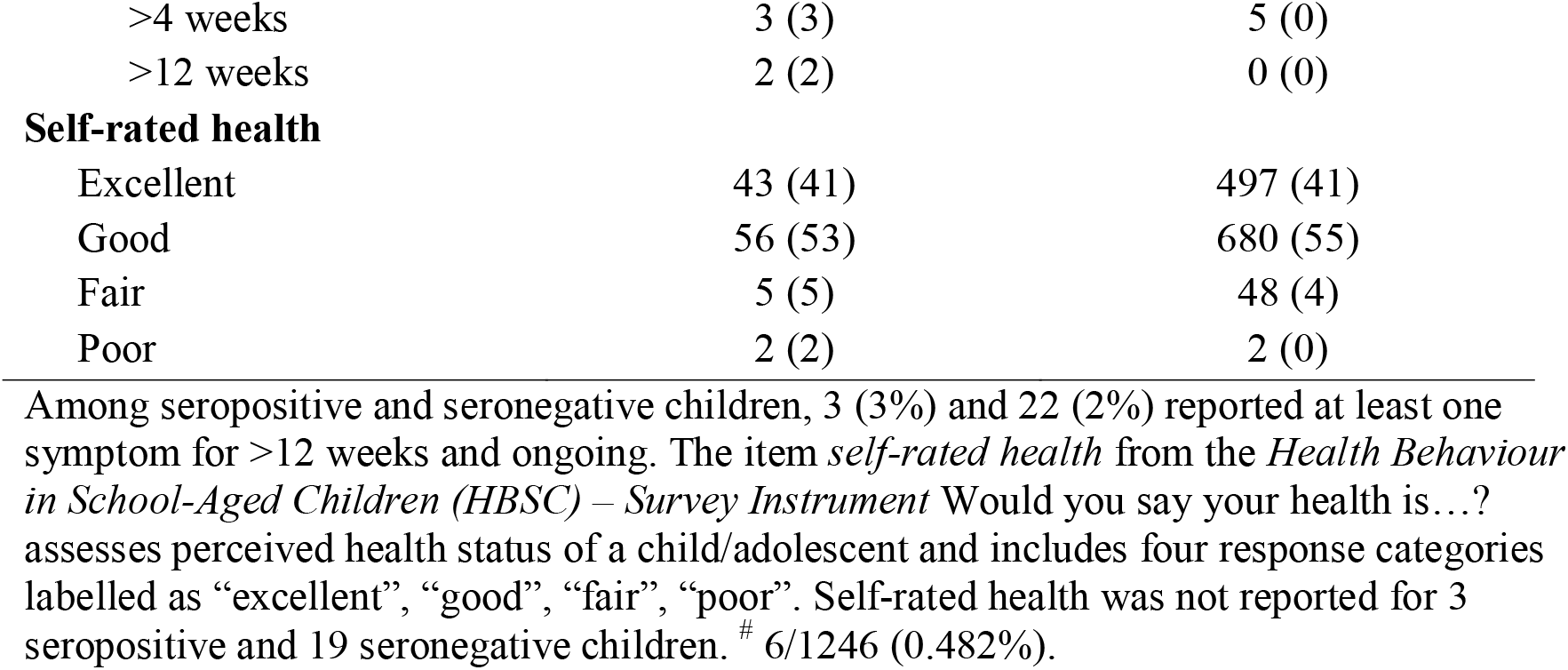
Participant characteristics, symptoms and self-rated health among seropositive and seronegative children.

|  | No. (%) |  |
| --- | --- | --- |
|  | Seropositive<br>(n=109) | Seronegative<br>(n=1246) |
| **Sex (female)** | 58 (53) | 669 (54) |
| **Age (years)** |  |  |
| 6-11 | 66 (61) | 703 (56) |
| 12-16 | 43 (39) | 543 (44) |
| **Persistent symptoms** |  |  |
| At least one symptom |  |  |
| >4 weeks | 10 (9) | 121 (10) |
| >12 weeks | 4 (4) | 28 (2) |
| 1-2 symptoms |  |  |
| >4 weeks | 2 (2) | 90 (7) |
| >12 weeks | 1 (1) | 22 (2) |
| 3 or more symptoms |  |  |
| >4 weeks | 8 (7) | 31 (2) |
| >12 weeks | 3 (3) | 6 (0)<sup>#</sup> |
| **Frequently reported symptoms** |  |  |
| Tiredness |  |  |
| >4 weeks | 7 (6) | 51 (4) |
| >12 weeks | 3 (3) | 10 (1) |
| Headache |  |  |
| >4 weeks | 5 (5) | 39 (3) |
| >12 weeks | 0 | 10 (1) |
| Congested or runny nose |  |  |
| >4 weeks | 3 (3) | 40 (3) |
| >12 weeks | 1 (1) | 3 (0) |
| Stomachache |  |  |
| >4 weeks | 3 (3) | 18 (1) |
| >12 weeks | 1 (1) | 3 (0) |
| Sleep disturbances |  |  |
| >4 weeks | 3 (3) | 14 (1) |
| >12 weeks | 0 | 6 (0) |
| Cough |  |  |
| >4 weeks | 2 (2) | 15 (1) |
| >12 weeks | 0 | 2 (0) |
| Difficulties concentrating |  |  |
| >4 weeks | 2 (2) | 12 (1) |
| >12 weeks | 2 (2) | 8 (1) |
| Increased sleep need |  |  |
| >4 weeks | 3 (3) | 5 (0) |
| >12 weeks | 2 (2) | 0 (0) |
| **Self-rated health** |  |  |
| Excellent | 43 (41) | 497 (41) |
| Good | 56 (53) | 680 (55) |
| Fair | 5 (5) | 48 (4) |
| Poor | 2 (2) | 2 (0) |

Among seropositive and seronegative children, 3 (3%) and 22 (2%) reported at least one symptom for >12 weeks and ongoing. The item *self-rated health* from the *Health Behaviour in School-Aged Children (HBSC) – Survey Instrument* Would you say your health is...? assesses perceived health status of a child/adolescent and includes four response categories labelled as “excellent”, “good”, “fair”, “poor”. Self-rated health was not reported for 3 seropositive and 19 seronegative children. # 6/1246 (0.482%).

## DISCUSSION

This study suggests a low prevalence of symptoms compatible with long COVID in a randomly selected population-based cohort of children followed over 6 months after serological testing. Seropositive children, with mostly asymptomatic SARS-CoV-2 infection, did not report these symptoms lasting for longer than 12 weeks more frequently than seronegative children.

While even severe forms of long COVID in children exist^4^, the estimates on prevalence range from 95% of children reporting symptoms within 8 months of follow-up^3^, to 1.8% of schoolchildren at 2 months in a large surveillance study^2^ or to full recovery in all children with predominantly mild disease^5^. Initial SARS-CoV-2 infection severity, different methodological approaches (clinical assessment *versus* self-report), definition of cases (diagnosed *versus* suspected cases), variable follow-up times, and prevalence of pre-existing chronic health conditions likely contribute to the variability of long COVID reported in children. Longitudinal data on large population-based samples are needed to better understand its potential impact on health-related quality of life and activities in daily living including going to school.

Strengths of our study include the large, representative, randomly selected sample of school children and inclusion of a population-based seronegative control group that could be ensured thanks to the longitudinal design. Limitations include the relatively small number of seropositive children, possible misclassification of some false seropositive or seronegative children, potential recall bias, parental report of child’s symptoms, and lack of information on symptom severity.

## Data Availability

All data are available on reasonable request.

