## Supplement Table 1-2 for "Long-term symptoms after SARS-CoV-2 infection in school children: population-based cohort with 6-months follow-up"

**Supplemental Table 1** Distribution of pre-existing chronic health conditions among seropositive and seronegative children

|  | **No. (%)** | |
| --- | --- | --- |
|  | **Seropositive**  (n=109) | **Seronegative**  (n=1246) |
| **Number of chronic health conditions** |  |  |
| 0 | 89 (82) | 894 (72) |
| 1 | 17 (16) | 245 (20) |
| 2 | 2 (2) | 70 (6) |
| 3 | 0 | 30 (2) |
| 4 or more | 1 (1) | 5 (0) |
| **Most frequent chronic health conditions** |  |  |
| Hay fever | 8 (7) | 185 (15) |
| Allergies | 6 (6) | 79 (6) |
| Eczema or neurodermatitis | 5 (5) | 70 (6) |
| Attention deficit/hyperactivity disorder | 2 (2) | 57 (5) |
| Asthma | 1 (1) | 55 (4) |

**Supplemental Table 2** Participant characteristics, symptoms and self-rated health among seropositive and seronegative children: children with chronic health conditions excluded

|  | **No. (%)** | |
| --- | --- | --- |
|  | **Seropositive**  (n=89) | **Seronegative**  (n=891) |
| **Sex (female)** | 48 (54) | 484 (54) |
| **Age (years)** |  |  |
| 6-11 | 56 (63) | 526 (59) |
| 12-16 | 33 (37) | 365 (41) |
| **Persistent symptoms** |  |  |
| 1-2 symptoms  >4 weeks  >12 weeks | 1 (1)  0 (0) | 64 (7)  12 (1) |
| 3 or more symptoms  >4 weeks  >12 weeks | 6 (7)  2 (2) | 16 (2)  3 (0) |
| **Frequently reported symptoms** |  |  |
| Tiredness  >4 weeks  >12 weeks | 5 (6)  2 (2) | 33 (4)  8 (1) |
| Headache  >4 weeks  >12 weeks | 4 (4)  0 (0) | 24 (3)  3 (0) |
| Congested or runny nose  >4 weeks  >12 weeks | 2 (2)  1 (1) | 24 (3)  1 (0) |
| Stomachache  >4 weeks  >12 weeks | 3 (3)  1 (1) | 7 (1)  0 (0) |
| Sleep disturbances  >4 weeks  >12 weeks | 3 (3)  0 (0) | 9 (1)  3 (0) |
| Cough  >4 weeks  >12 weeks | 2 (2)  0 (0) | 10 (1)  1 (0) |
| Difficulties concentrating  >4 weeks  >12 weeks | 1 (1)  1 (1) | 6 (1)  3 (0) |
| Increased sleep need  >4 weeks  >12 weeks | 1 (1)  1 (1) | 4 (0)  0 (0) |
| **Self-rated health** |  |  |
| Excellent | 38 (44) | 372 (43) |
| Good | 43 (50) | 465 (53) |
| Fair | 4 (5) | 37 (4) |
| Poor | 1 (1) | 1 (0) |

Among seropositive and seronegative children, 2 (2%) and 13 (1%) reported at least one symptom for >12 weeks and symptoms. The item *self-rated health* from the *Health Behaviour in School-Aged Children (HBSC) – Survey Instrument* “Would you say your health is…?” assesses perceived health status of a child/adolescent and includes four response categories labelled as “excellent”, “good”, “fair”, “poor”. Self-rated health was not reported for 3 seropositive and 16 seronegative children.
